# Improving Smoking History Documentation to Facilitate Lung Cancer Screening Utilization

**DOI:** 10.64898/2025.12.24.25342974

**Authors:** Yannan Lin, Ruiwen Ding, Seyed Mohammad Hossein Tabatabaei, Haley I Tupper, Drew Moghanaki, Brett H. Schussel, Denise R. Aberle, William Hsu, Ashley Elizabeth Prosper

## Abstract

**Objectives:** Lung cancer screening (LCS) is the only screening test incorporating behavioral risk factors into eligibility determination. However, collecting necessary smoking history data has been challenging, limiting screening uptake. In this study, we evaluated how a program coordinator’s detailed shared decision-making (SDM) impacted smoking data reliability.

**Methods:** Patients who underwent a baseline screening low-dose CT between July 31, 2013, and August 25, 2023, were stratified into pre- and post-intervention cohorts. The intervention was a comprehensive pre-CT smoking history assessment with SDM by an LCS program coordinator, implemented on July 31, 2017. We compared the completeness and concordance of smoking history data between clinician and patient self-report.

**Results:** Among 3795 patients, 670 (18%) were pre- and 3125 (82%) were post-intervention. Having a coordinator reduced missing smoking data (p<0.001), but did not eliminate it. Both groups showed high concordance between clinician-documented and self-reported smoking status (pre: kappa=0.84, 95% confidence interval [CI] 0.79-0.89; post: kappa=0.84, 95% CI 0.83-0.86). Correlations strengthened for smoking duration (rho=0.71 vs. 0.65, p=0.026) and years since quitting (rho=0.83 vs. 0.80, p=0.21) after involving a coordinator. Correlations for smoking intensity and pack years remained fair (rho<0.6). LCS eligibility based on self-reported smoking history increased from 46.0% (308/670) pre- to 64.1% (2003/3125) post-intervention, below the 100% eligibility using clinician-documented history.

**Conclusions:** Smoking data reliability improved after a dedicated LCS program coordinator implemented a smoking history assessment. Meanwhile, challenges remained with the ascertainment of total pack-years. Detailed probing and patient education may be insufficient to overcome challenges in assessing smoking intensity.

## Introduction

Lung cancer is the leading cause of cancer mortality in the United States (US) in both sexes.^1^ Lung cancer screening (LCS) with annual low-dose computed tomography (LDCT) reduces mortality by 20%-30%.^2, 3^ Annual LDCT screening has been recommended by the United States Preventive Services Task Force (USPSTF) since 2013 for high-risk individuals, with the latest 2021 update expanding eligibility to individuals aged 50 to 80 years with a minimum 20-pack-year smoking history who currently smoke or have quit smoking within the past 15 years.^4^ However, LCS faces crucial challenges in adoption, with only 16% of eligible individuals screened in 2022.^5^ Low participation in LCS misses opportunities for earlier detection among eligible individuals.

Factors associated with LCS non-participation are multi-faceted, involving patients, providers, and the healthcare system.^6, 7^ At the provider level, challenges in identifying eligible patients from electronic medical records (EMRs) are difficult to overcome. The determination of population-wide LCS eligibility is impeded by inadequate documentation of smoking history in the EMR, especially the pack-years information. An analysis of Veterans Health Administration (VHA) hospital records published in 2016 revealed that the smoking status or pack-year history could not be determined from the EMR for 39% (36,555 out of 93,033) of primary care patients.^8^ A separate study that surveyed 110 Federally Qualified Health Centers demonstrated only 54% of patient records had documentation of pack-year history necessary for LCS eligibility, and only 29% of providers with access to these data considered it reliably accurate for patient care decisions.^9^

Given the complexities of ascertaining LCS eligibility, which requires a complex coordination of care across multiple levels, including documentation of a shared decision-making discussion.^10^ Our institution employed a program coordinator to help with the requirements. The coordinator conducted SDM visits when documentation was lacking, and attempted each time to confirm an accurate smoking history for the medical record. Our institution’s program also collects smoking history data via a pre-screening questionnaire and has access to patient self-reported data if the data is missing from the EMR. The availability of two distinct data sources for smoking history provides an opportunity to assess the impact of the LCS program coordinator on smoking data documentation to facilitate LCS.

To our knowledge, we are among the first to leverage these data sources to evaluate whether the presence of a program coordinator improved the quality of smoking history documentation, including the notoriously challenging pack years data. We hypothesized that having a program coordinator improved the correlation between self-reported and clinician-documented smoking histories and reduced missing values. We also explored factors contributing to discrepancies between these data sources and their implications for our LCS program.

## Methods

### Patient enrollment

Internal Board Review (IRB) approval was obtained at our institution for conducting this retrospective cohort study. Informed consent was waived because the risk to patients was minimal. Patients who underwent their baseline LDCT screening at our institution between July 31, 2013, and August 25, 2023, and who met the USPSTF LCS eligibility criteria at the time of baseline screening, as determined by clinician-documented smoking history, were included. This study followed the Strengthening the Reporting of Observational Studies in Epidemiology (STROBE) reporting guideline for cohort studies.

### Patient subgroups

A nurse practitioner coordinator joined the LCS program at our institution on July 31, 2017. Patients were categorized into two groups: those enrolled without (between July 31, 2013 and July 30, 2017) and with (on and after July 31, 2017) a program coordinator.

### Data collection

Patient information at the baseline screen was extracted from the EMR system, including age, primary insurance, clinician-documented smoking history, and Lung CT Screening Reporting & Data System (Lung-RADS) score. Clinician-documented smoking history, including smoking status, smoking intensity (i.e., the average number of cigarettes per day), smoking duration (i.e., years smoked, the number of years an individual has smoked cigarettes), and years since quitting (i.e., the number of years an individual has quit smoking cigarettes), was manually extracted from clinical notes. Pack-years were calculated using the following formula: (average number of cigarettes per day/20) × years smoked. Shortly after the SDM visit (typically within two weeks), patients would report their smoking history in the questionnaire during their scheduled baseline screening LDCT appointment. The patient’s self-reported smoking history was obtained from this questionnaire routinely administered to all patients undergoing LCS prior to the imaging examination and stored as a discrete series of the screening examination in our picture archiving and communication system (see **Table S1** in the Supplement). Relevant data fields and options included smoking status, starting age, quitting age, years smoked, and average number of cigarettes/day. The questionnaire also collected other variables required by the PLCOm2012 risk model, including sex, race, and ethnicity. A manual chart review addressed missing race and ethnicity if these were not recorded on the self-report questionnaire.

### Interrater reliability of extracted smoking data from EMR

Rules were first developed by one co-author (Y.L.) to aid in extracting quantitative smoking information (i.e., intensity, duration, years since quitting, and pack-years) from the chart, where data from the SDM visit note was prioritized, if available, followed by the primary care physician visit and other notes closest to the baseline LDCT screening date. Fifty patients were randomly selected to validate the rules. Three co-authors (Y.L., R.D., and S.M.H.T.) first extracted the smoking history for 25 patients, followed by three group meetings (i.e., Y.L. and R.D., Y.L., and S.M.H.T, and all three co-authors) to address questions and discuss considerable discrepancies to refine the data abstraction rules (e.g., the mean was used if a range was mentioned in a note, see **Supplemental Methods**). The remaining 25 patients served as the validation subjects, where the three raters agreed on smoking intensity, duration, years since quitting, and pack-years (two-way mixed effects model, single rater absolute intraclass correlation coefficient between 0.95 and 0.98).^11^ After the validation, the clinician-documented smoking history was extracted by one of the authors (Y.L.), who has seven years of experience with data collection from EMR, following the refined rules.

### Statistical analysis

Data were summarized for groups with (post-intervention) and without (pre-intervention) a program coordinator. Smoking data completeness and consistency were evaluated among patients with both self-reported and clinician-documented smoking history. Cohen’s Kappa, with a 95% confidence interval (CI), assessed the agreement between self-reported and clinician-documented smoking status. The interpretation of Cohen’s Kappa values was the following: ≤ 0 indicating no agreement and 0.01–0.20 as none to slight, 0.21–0.40 as fair, 0.41– 0.60 as moderate, 0.61–0.80 as substantial, and 0.81–1.00 as almost perfect agreement.^12^ When comparing categorical variables, including smoking status, data missingness, and proportion eligible for LCS, McNemar’s test was used for paired data, and Fisher’s exact or Chi-square test was used for unpaired data. For continuous smoking variables (i.e., intensity, duration, years since quitting, and pack-years), Spearman’s correlation coefficient was used to determine the monotonic association between the two smoking measures. The correlation coefficients were interpreted as poor if less than 0.3, fair if 0.3 to <0.6, moderately strong if 0.6 to <0.8, and very strong if at least 0.8.^13^ Fisher’s z transformation was applied to compare two correlation coefficients. The Wilcoxon rank sum test assessed the difference between self-reported and clinician-documented continuous smoking measures. Years since quitting were compared among former smokers only. Scatter plots between self-reported and clinician-documented continuous smoking measures were generated. For the categorical smoking status variable, ten patients were randomly selected and their medical records were reviewed to identify potential reasons for data inconsistency between the two sources. For other continuous smoking variables, patients with the top ten greatest differences (i.e., five where clinician-documented values exceeded self-reported values and five where clinician-documented values were lower) were examined to determine possible reasons for inconsistency. Patients with missing smoking history data were compared to complete records for each data source separately (see **Supplemental Methods and Results** and **Tables S2 and S3**). Data analysis was performed using R (version 3.6.1, R Core Team, Vienna, Austria) and Python (version 3.7.3, Python Software Foundation). Two-sided p< .05 was considered significant.

### Data availability

The data underlying this study cannot be shared publicly to protect the privacy of study participants.

## Results

### Patient characteristics

A total of 3795 patients who underwent a baseline LDCT screen at our institution between July 31, 2013, and August 25, 2023, met the appropriate USPSTF eligibility criteria determined by clinician-documented smoking history. Median (IQR) age was 64 (59-69) years; 1445 patients (38.1%) were female; 2350 (61.9%) were male; 19 (0.5%) American Indian or Alaska Native, 311 (8.2%), Asian; 232 (6.1%), Black; 239 (6.3%), Hispanic; 2792 (73.6%), White; and 30 (0.8%), other race and ethnicity. Most patients (2390 [63.0%]) had private or commercial insurance. There were 670 and 3125 patients in the groups without and with a program coordinator, respectively. **Table** 4 in the Supplement summarizes patient characteristics in the two groups. Sex and race/ethnicity distributions were similar between those managed before and after the hiring of an LCS program coordinator. In the group with a program coordinator, age was lower (64 vs. 66, p<0.05), and the proportion of those with commercial or private insurance was higher (66% vs. 49%, p<0.05). **Table 1** details the comparison between self-reported and clinician-documented smoking history between the two groups. In the group without a program coordinator, clinician-documented mean smoking intensity (average cigarettes smoked/day) (23.8 vs. 19.4, p<0.001) and pack years (46.1 vs. 37.8, p<0.001) were significantly higher than patient self-report. The findings for smoking intensity (21.4 vs. 17.7, p<0.001) and pack years (39.1 vs. 32.6, p<0.001) were similar in the group with a program coordinator. In addition, the program coordinator group had a slightly higher proportion of clinician-documented current smokers than the self-reported data (42.5% vs. 41.6%, p<0.001). The clinician documented years since quitting was significantly lower than the self-reported information in the group with a program coordinator (6.6 vs. 8.0, p=0.002).

**Table 1.** Comparison of self-reported and clinician-documented smoking history.

| Variable | Without program coordinator (n=670) |  |  | With program coordinator (n=3125) |  |  |
| --- | --- | --- | --- | --- | --- | --- |
|  | Self-report | EMR | p-value | Self-report | EMR | p-value |
| Smoking status n (%) |  |  | 0.47 |  |  | <0.001 |
| Current | 240 (48.6) | 236 (47.8) |  | 1277 (41.6) | 1306 (42.5) |  |
| Former | 254 (51.4) | 258 (52.2) |  | 1796 (58.4) | 1767 (57.5) |  |
| Missing (%) | 176 (26.3) | 0 (0) |  | 52 (1.7) | 0 (0) |  |
| Average number of cigarettes/day (intensity) mean (sd) | 19.4 (9.9) | 23.8 (9.8) | <0.001 | 17.7 (8.4) | 21.4 (8.5) | <0.001 |
| Missing (%) | 81 (12.1) | 54 (8.1) |  | 50 (1.6) | 65 (2.1) |  |
| Years smoked (duration) mean (sd) | 40.3 (11.0) | 40.5 (9.4) | 0.52 | 37.1 (10.9) | 37.5 (9.8) | 0.95 |
| Missing (%) | 180 (26.9) | 53 (7.9) |  | 63 (2.0) | 61 (2.0) |  |
| Years since quitting mean (sd) <sup>s</sup> | 7.3 (7.1) | 6.1 (4.6) | 0.18 | 8.0 (7.2) | 6.6 (4.7) | 0.002 |
| Missing (%) | 7 (2.8) | 0 (0) |  | 41 (2.3) | 0 (0) |  |
| Pack-years mean (sd) | 37.8 (23.6) | 46.1 (18.2) | <0.001 | 32.6 (18.4) | 39.1 (17.4) | <0.001 |
| Missing (%) | 186 (27.8) | 0 (0) |  | 103 (3.3) | 0 (0) |  |

Notes: <sup>a</sup> Without a program coordinator, there were 254 former smokers in self-reported data and 359 in clinician-documented data. With a coordinator, these numbers were 1796 and 1801, respectively.
Abbreviations: EMR: electronic medical record; sd: standard deviation.

### Smoking data completeness

Patients undergoing LCS before versus after the introduction of an LCS program coordinator had complete clinician-documented smoking status, years since quitting, and pack years data since these data elements are required to determine LCS eligibility at our institution (**Table 2**). For self-reported (all smoking variables) and clinician-documented (intensity and duration) smoking history, there was a significant reduction in the proportion of missing values for each variable in the group with a program coordinator as opposed to the group with one (p<0.001). Combining both patient self-report and clinician-documented smoking data, complete documentation of smoking history for all patients was accomplished.

**Table 2.** Comparison of proportions of missing values in smoking history data.

| Variable | Without a<br>program<br>coordinator,<br>n=670 | With a<br>program<br>coordinator,<br>n=3125 | p-value |
| --- | --- | --- | --- |
| Self report |  |  |  |
| Smoking status | 176 (26.3) | 52 (1.7) | <0.001 |
| Average number of cigarettes/day (intensity) | 81 (12.1) | 50 (1.6) | <0.001 |
| Years smoked (duration) | 180 (26.9) | 63 (2.0) | <0.001 |
| Years since quitting<sup>a</sup> | 7 (2.8) | 41 (2.3) | <0.001 |
| Pack-years | 186 (27.8) | 103 (3.3) | <0.001 |
| EMR |  |  |  |
| Smoking status | 0 (0) | 0 (0) | NA |
| Average number of cigarettes/day (intensity) | 54 (8.1) | 65 (2.1) | <0.001 |
| Years smoked (duration) | 53 (7.9) | 61 (2.0) | <0.001 |
| Years since quitting<sup>a</sup> | 0 (0) | 0 (0) | NA |
| Pack-years | 0 (0) | 0 (0) | NA |

Notes: <sup>a</sup> Without a program coordinator, there were 254 former smokers in self-reported data and 359 in clinician-documented data. With a coordinator, these numbers were 1796 and 1801, respectively.
Abbreviations: EMR: electronic medical record; NA: not available.

### Smoking data consistency

In **Table 3**, the agreement between self-reported and clinician-documented smoking status was almost perfect regardless of the presence of a program coordinator (without: κ=0.84, 95% CI 0.79-0.89; with: κ=0.84, 95% CI 0.83-0.86). The correlation between self-reported and clinician-documented smoking intensity was fair in both groups, with an increase observed in the program coordinator group. However, the improved concordance did not reach statistical significance (without: ρ=0.49, with: ρ=0.55, p=0.08). Compared to the group without a program coordinator, the group with a program coordinator had a significantly higher correlation between self-reported and clinician-documented smoking duration (without: ρ=0.65, with: ρ=0.71, p=0.026). The agreement for smoking duration between the two data sources was moderately strong for both groups. Among former smokers, the correlation between the two groups was very strong, with a higher correlation in the program coordinator group, which did not achieve statistical significance (without: ρ=0.80, with: ρ=0.83, p=0.21). Pack-years showed a higher correlation in the program coordinator group (without: ρ=0.40, with: ρ=0.55, p<0.001), but overall, the correlation remained fair. **Figure S1** in the Supplement delineates the association between continuous self-reported and clinician-documented smoking variables.

**Table 3.** Agreement and correlation between self-reported and clinician-documented smoking history.

| Variable | Without program coordinator |  | With program coordinator |  | p-value<sup>a</sup> |
| --- | --- | --- | --- | --- | --- |
| | n | $\kappa$ (95% CI) | n | $\kappa$ (95% CI) | |
| Smoking status | 494 | 0.84 (0.79-0.89) | 3073 | 0.84 (0.83-0.86) |  |
| | n | $\rho$ (p-value) | n | $\rho$ (p-value) | |
| Average number of cigarettes/day (intensity) | 550 | 0.49 (<0.001) | 3012 | 0.55 (<0.001) | 0.08 |
| Years smoked (duration) | 456 | 0.65 (<0.001) | 3003 | 0.71 (<0.001) | 0.026 |
| Years since quitting | 229 | 0.80 (<0.001) | 1638 | 0.83 (<0.001) | 0.21 |
| Pack-years | 484 | 0.39 (<0.001) | 3022 | 0.55 (<0.001) | <0.001 |

Notes: <sup>a</sup> Fisher's z transformation. $\kappa$ : Cohen's kappa. $\rho$ : Spearman's rank correlation coefficient.
Abbreviations: CI: confidence interval.

**Table 4.**
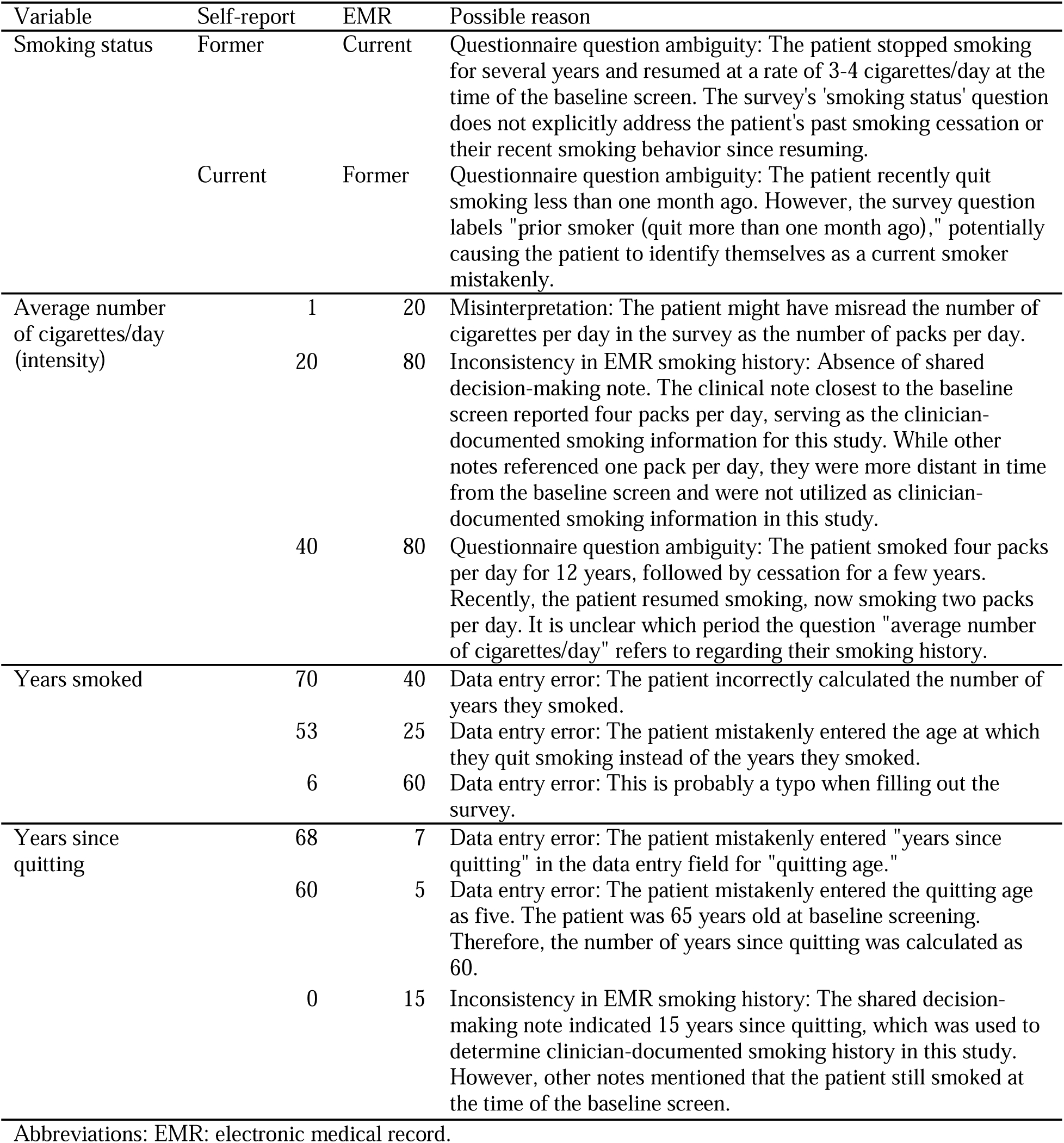
Examples of differences in smoking history documentation.

| Variable | Self-report | EMR | Possible reason |
| --- | --- | --- | --- |
| Smoking status | Former | Current | Questionnaire question ambiguity: The patient stopped smoking for several years and resumed at a rate of 3-4 cigarettes/day at the time of the baseline screen. The survey's 'smoking status' question does not explicitly address the patient's past smoking cessation or their recent smoking behavior since resuming. |
|  | Current | Former | Questionnaire question ambiguity: The patient recently quit smoking less than one month ago. However, the survey question labels "prior smoker (quit more than one month ago)," potentially causing the patient to identify themselves as a current smoker mistakenly. |
| Average number of cigarettes/day (intensity) | 1 | 20 | Misinterpretation: The patient might have misread the number of cigarettes per day in the survey as the number of packs per day. |
|  | 20 | 80 | Inconsistency in EMR smoking history: Absence of shared decision-making note. The clinical note closest to the baseline screen reported four packs per day, serving as the clinician-documented smoking information for this study. While other notes referenced one pack per day, they were more distant in time from the baseline screen and were not utilized as clinician-documented smoking information in this study. |
|  | 40 | 80 | Questionnaire question ambiguity: The patient smoked four packs per day for 12 years, followed by cessation for a few years. Recently, the patient resumed smoking, now smoking two packs per day. It is unclear which period the question "average number of cigarettes/day" refers to regarding their smoking history. |
| Years smoked | 70 | 40 | Data entry error: The patient incorrectly calculated the number of years they smoked. |
|  | 53 | 25 | Data entry error: The patient mistakenly entered the age at which they quit smoking instead of the years they smoked. |
|  | 6 | 60 | Data entry error: This is probably a typo when filling out the survey. |
| Years since quitting | 68 | 7 | Data entry error: The patient mistakenly entered "years since quitting" in the data entry field for "quitting age." |
|  | 60 | 5 | Data entry error: The patient mistakenly entered the quitting age as five. The patient was 65 years old at baseline screening. Therefore, the number of years since quitting was calculated as 60. |
|  | 0 | 15 | Inconsistency in EMR smoking history: The shared decision-making note indicated 15 years since quitting, which was used to determine clinician-documented smoking history in this study. However, other notes mentioned that the patient still smoked at the time of the baseline screen. |

Abbreviations: EMR: electronic medical record.

### Self-reported smoking history for LCS eligibility

**Table S5** in the Supplement demonstrates the determination of LCS eligibility using patient self-reported smoking history according to the USPSTF guidelines. In the group with a program coordinator, compared to the group without a program coordinator, the proportion of patients eligible for LCS by the USPSTF guidelines increased from 46.0% (308/670) to 64.1% (2003/3125), and the proportion of patients with undetermined eligibility due to missing values in smoking history decreased from 28.7% (192/670) to 4.4% (136/3125); however, the proportion of ineligible patients as determined using patient self-reported smoking data increased from 25.4% (170/670) to 31.6% (987/3125).

### Examples of data discrepancies and potential reasons

After reviewing ten patients with the greatest differences between the two data sources for each variable, typical scenarios of differences between the two smoking data sources were summarized in **Table 4**. Common reasons for inconsistencies between patient self-reported and clinician-documented smoking history may be attributed to either source, such as 1) questionnaire question ambiguity, 2) misinterpretation of questionnaire questions, 3) questionnaire data entry error, and 4) EMR smoking data inconsistency.

## Discussion

Documentation of smoking status and years since quit achieved high agreement and correlation between self-report and clinician documentation regardless of intervention by a LCS program coordinator. The group with a program coordinator had a higher correlation between self-reported and clinician-documented smoking duration, achieving a moderately strong correlation. Pack-year documentation benefited from the presence of an LCS program coordinator, however, the correlation remained fair even with the coordinator’s presence. This is due to the poor correlation in smoking intensity between the two data sources (pack-years = intensity × duration), highlighting the challenge of consistently recalling the average number of cigarettes per day among smokers. Having a program coordinator eliminated missing values in smoking history. By combining both sources of data, a complete smoking history can be ensured.

The regular responsibilities of an LCS program coordinator involve coordinating LCS-related activities and improving annual adherence to lung screening.^10, 14, 15^ Our findings underscore the value of tasking a program coordinator with the review of smoking history with patients to document more consistent information in the EMR. Our study demonstrates improved correlation between patient self-reported and clinician-documented smoking history after the introduction of an LCS program coordinator. A higher correlation may stem from the framework (i.e., a comprehensive pre-CT smoking history assessment with SDM) that our LCS program coordinator establishes during an SDM discussion which may improve patient recall.

Similar to our findings, previous work has reported high agreement between self-reported and EMR-reported smoking status in a non-LCS cohort (κ=0.85, 95% CI 0.75–0.96).^16^ While we showed that a program coordinator improves the completeness of pack-year data, our findings confirm previous findings that highlight the difficulty of achieving accurate smoking history documentation.^8, 9^ This ongoing challenge impedes the expansion of LCS programs in the US. Nationwide initiatives are using survey-based eligibility determination tools to boost LCS participation rates. Available LCS eligibility determination aid tools^17, 18^ often employ similar survey questions as our questionnaire. Patients may mistakenly answer survey questions and receive an ineligible LCS result despite being eligible. To address this, we recommend adding a note on the result page advising patients to “consult their healthcare provider for further evaluation of LCS eligibility,” as smoking history documentation may be more accurate when ascertained by a clinician. Our findings highlight the need to reassess the process for collecting total pack-years, as current questions developed by medical professionals may not be the most practical for patients. Even at current LCS uptake rates, reliance on a coordinator to document pack-years is challenging, due to cost and time constraints. As rates of LCS uptake increase, this model will likely prove to be unsustainable. Simplifying the questions for self-reporting would improve accessibility, increase completion rates, and ensure broader participation in LCS.

While total pack-years remain a conventional measure of cigarette smoke exposure^19^, research suggests that replacing the 20 total pack-years criterion with a 20-year smoking duration criterion could increase the proportion of at-risk patients eligible for LCS while reducing racial disparities in eligibility.^20^ While the LCS eligibility criteria have undergone revision, some individuals go on to develop lung cancer before meeting criteria, as many as 46–54% of lung cancer cases in one cohort.^21^ Compared to the fair correlation for pack-years, a moderately strong correlation between self-report and clinician documentation of smoking duration suggests that it may be an easier-to-implement metric for determining LCS eligibility. In addition, the National Committee for Quality Assurance has begun the development of HEDIS^®^ quality measures for LCS and Tobacco Use Screening and Cessation^22^, where the quality of smoking history data is essential for accurately measuring screening uptake and tobacco use. These highlight the critical role of reliable smoking documentation beyond determining LCS eligibility, that it is also critical in optimizing LCS guidelines, promoting equitable access, and supporting quality measures in lung cancer prevention.

Gaps persist in the smoking history documentation between clinician and patient-derived sources. The group with a program coordinator had minimal missing values in smoking history documentation. However, it remains to be investigated why approximately one-third of patients are deemed ineligible for screening based solely on self-reported smoking history. This discrepancy could be partly attributed to potential issues identified with completing the questionnaire, as outlined in **Table 4**, or patients underreporting their smoking habits due to stigma.^23^ Possible solutions to address these issues may include assigning personnel to verify the smoking history section upon questionnaire completion to capture and correct obvious data entry errors, clarifying that the average number of cigarettes per day refers to lifetime rather than recent smoking, and revisiting the calculation-error-prone “years smoked” field as it can be derived from existing data elements (age at baseline screen minus starting age for current smokers, and quitting age minus starting age for former smokers). The findings of this study have been shared with our LCS program staff to aid in revising the questionnaire.

Besides improving self-reported smoking data, efforts are necessary to improve the EMR documentation of smoking history. Several issues were noticed during the manual chart review to extract clinician-documented smoking history, such as missing values (missing intensity and duration in **Table 4**), outdated data, discrepancies across different physicians, etc. Approaches to improve EMR smoking data quality may include applying data collection and analysis strategies that reflect changes in smoking habits over time and post-extraction processing, such as applying multiple imputation and pattern matching algorithms.^24, 25^

## Limitations

This study examines agreement between two data sources for smoking information and the coordinator’s role in improving consistency and completeness. However, it does not directly determine the optimal source for obtaining the most accurate smoking history information. In addition to the program coordinator, changes in smoking history documentation could be attributed to improvements in EMR systems and greater awareness of the importance of accurate documentation over time. Self-reported smoking history may be subject to recall bias.^26^ Notably, recall bias may be less prominent for patients in the program coordinator group, as they have undergone a review of their smoking history with a clinician during an SDM or other visit before completing the questionnaire. Additionally, we established rules to extract quantitative smoking information from EMR. However, our rules may only address certain inconsistencies in longitudinal documentation of smoking history in the medical records. Further research is warranted to fully understand the clinician-documented longitudinal smoking history discrepancies within our cohort. Moreover, only limited possible reasons for smoking data inconsistencies were identified, which needed to be further verified with the patients. Our study highlights the completeness and consistency of smoking history between the two data sources. Other aspects of data quality assessment, such as accuracy and timeliness, remain to be investigated. A cost-benefit analysis comparing the two data sources would be helpful to guide decisions on resource allocation and the most efficient strategies for improving smoking data quality and screening outreach.

## Conclusions

Hiring a program coordinator improved smoking data quality, but was unable to overcome historical challenges in ascertaining accurate pack-year data required to confirm LCS eligibility. Ambiguous questionnaire items for self-reporting and discrepancies in clinician documentation of smoking history need to be addressed. We recommend engaging patients in the completion of current questionnaires to improve pack-year ascertainment and in turn, improve the efficiency of LCS eligibility determination.

## Author contributions

Yannan Lin: Conceptualization; Data curation; Formal analysis; Investigation; Methodology; Project administration; Resources; Software; Validation; Visualization; Writing—original draft; Writing—review & editing

Ruiwen Ding: Conceptualization; Data curation; Investigation; Methodology; Validation; Writing—review & editing

Seyed Mohammad Hossein Tabatabaei: Conceptualization; Data curation; Investigation; Methodology; Validation; Writing—review & editing

Haley I Tupper: Conceptualization; Investigation; Visualization; Writing—original draft; Writing—review & editing

Drew Moghanaki: Conceptualization; Investigation; Visualization; Writing—original draft; Writing—review & editing

Brett H. Schussel: Conceptualization; Investigation; Resources; Visualization; Writing—original draft; Writing—review & editing

Denise R. Aberle: Conceptualization; Funding acquisition; Resources; Supervision; Writing—review & editing

William Hsu: Conceptualization; Funding acquisition; Resources; Supervision; Writing—review & editing

Ashley Elizabeth Prosper: Conceptualization; Funding acquisition; Resources; Supervision; Writing—review & editing

## Statements and Declarations

Yannan Lin reports financial support was provided by NIH National Institute of Biomedical Imaging and Bioengineering. Yannan Lin reports financial support was provided by the Agency for Healthcare Research and Quality. Ruiwen Ding reports financial support was provided by the V Foundation. Ashley Elizabeth Prosper reports financial support was provided by the NIH National Cancer Institute. Drew Moghanaki reports financial support from the Stanley Iezman and Nancy Stark Endowment for Thoracic Radiation Oncology Research. Denise R. Aberle reports financial support was provided by NIH National Cancer Institute. William Hsu reports financial support was provided by the V Foundation. William Hsu reports financial support was provided by the NIH National Institute of Biomedical Imaging and Bioengineering. William Hsu reports financial support was provided by the NIH National Cancer Institute. William Hsu reports financial support was provided by the National Science Foundation. William Hsu reports financial support was provided by the Agency for Healthcare Research and Quality. Drew Moghanaki reports a relationship with LungLife AI that includes: stock options. Denise R. Aberle reports a relationship with the Kaiser Foundation Research Institute (Patient-Centered Outcomes Research Institute) that includes funding grants. Denise R. Aberle reports a relationship with DECAMP 1 PLUS (Janssen Pharmaceuticals) that includes: funding grants. Denise R. Aberle reports a relationship with V Foundation that includes: funding grants. Denise R. Aberle reports a relationship with LungLife AI that includes: funding grants. Denise R. Aberle reports a relationship with Liquid Diagnostics LL that includes: funding grants. Denise R. Aberle reports a relationship with EarlyDiagnostics that includes: funding grants. William Hsu reports a relationship with EarlyDiagnostics that includes: funding grants. William Hsu reports a relationship with the Radiological Society of North America that includes consulting or advisor.

## Ethical considerations

Internal Board Review (IRB) approval was obtained at our institution for conducting this retrospective cohort study.

## Consent to participate

Informed consent was waived because the risk to patients was minimal.

## Consent for publication

Informed consent was waived because the risk to patients was minimal. Publication plans are mentioned in the IRB.

## Data Availability

The data underlying this study cannot be shared publicly to protect the privacy of study participants.

## Data Availability

The study was performed with institutional review board approval and waiver of informed consent, and the data underlying this study cannot be shared publicly due to the privacy of individuals who participated in the study. Aggregated summaries without individual data were shared in the article.

